# Tongue swab-based Targeted Universal Tuberculosis Testing in people living with HIV in KwaZulu-Natal, South Africa

**DOI:** 10.64898/2026.04.17.26351084

**Authors:** Alaina M. Olson, Rachel C. Wood, Nsika Sithole, Indira Govender, Alison D. Grant, Theresa Smit, Anura David, Wendy Stevens, Lesley Scott, Paul K. Drain, Gerard A. Cangelosi, Adrienne E. Shapiro

## Abstract

**Background:** Targeted Universal Tuberculosis Testing (TUTT) may increase tuberculosis (TB) case detection by including people who are not actively seeking TB care but are at high risk of the disease.Non-invasive tongue swab (TS) testing may facilitate TUTT. We evaluated two TS testing protocols in people with HIV (PWH) tested irrespective of TB symptoms.

**Methods:** Study staff collected Copan FLOQSwab and Medline foam swab specimens, alongside urine and sputa, from PWH, most of whom were presenting for antiretroviral therapy initiation at primary healthcare clinics in KwaZulu-Natal, South Africa. FLOQSwabs were tested by sequence-specific magnetic capture (SSMaC) with qPCR (FLOQSwab-SSMaC). Foam swabs were tested by centrifuge-sedimentation and high-volume qPCR (foam-sedimentation). Urine lipoarabinomannan was detected using LF-LAM. The extended microbiological reference standard (eMRS) comprised any positive result on Xpert Ultra and/or liquid culture of sputum.

**Results:** We enrolled 251 participants (median age 34 years, 56% female, 67% with self-reported TB symptoms). Participants had a median CD4 count of 347 cells/µl, and 16% (40/251) had prior TB. FLOQSwab-SSMaC was 43% sensitive (13/30) and 100% specific (131/131) relative to eMRS. Foam-sedimentation was 47% (9/29) sensitive and 100% (176/176) specific. Sensitivity increased to 52% (FLOQSwab-SSMaC) and 50% (foam-sedimentation) when sputum Xpert Ultra Trace positive results were excluded from eMRS. TS was more sensitive than urine LAM, and both sample types were more sensitive when CD4 counts were below 200.

**Discussion:** TS testing detected about half of PWH with TB and outperformed urine LAM within this population, including among PWH with low CD4 counts.

**Summary:** TB testing approaches using tongue swabs (TS), combined with molecular methods, have the potential to identify people with TB in settings where sputum collection is not practical. People living with HIV are an at-risk group that could benefit from non-sputum based targeted TB testing programs.

## Introduction

To meet global goals for reducing tuberculosis (TB) transmission, morbidity and mortality, new strategies are needed for early detection (1-4). A major challenge is the underdiagnosis of TB disease, including among people living with HIV (PWH). TB can be difficult to diagnose in PWH despite their relatively high risk of developing the disease. *Mycobacterium tuberculosis* (MTB) is usually detected in sputum, a viscous material derived from patient airways. Sputum collection is challenging in community settings, and many patients cannot routinely produce sputum in any setting (1-4).

PWH are at elevated risk for TB disease, especially those who are not yet on antiretroviral therapy (ART). Active case finding, or screening for active TB among people at higher risk (such as PWH), could help improve the management of both diseases. TB detection in PWH and other vulnerable populations would be simplified by using a sampling method that is more easily and universally collectable than sputum. With its demonstrated ease of use (5-10), tongue swabbing (TS) is a promising non-sputum specimen for TB diagnosis that could greatly facilitate active TB case finding (8, 11).

The specificity of TS with qPCR read-outs to detect MTB DNA, relative to a sputum extended- and microbiological reference standard (e/MRS), is consistently high. However, sensitivity remains variable between 35% and 95% depending on population and methodology (7, 12-19). TS is very sensitive among symptomatic TB patients with high bacillary loads, but less so when applied to asymptomatic people and others with low bacillary loads (20). Nonetheless, in populations and settings where sputum collection is not feasible, TS could make it possible to find TB cases that would otherwise go undetected. The World Health Organization (WHO) announced its recommendation of the method in February 2026 (21).

This study evaluated the performance of TS testing in a population consisting of PWH most of whom were not actively seeking TB care when enrolled. In South Africa, all persons initiating HIV care are tested for TB using sputum testing according to guidelines of the National HIV Programme (22). To analyze the TS samples collected from such participants, we used laboratory methods that were explicitly designed for swab testing and exhibited high accuracy in past studies (12, 20) [newer, less labor-intensive near point-of-care (nPOC) methods (15, 19, 23, 24) were announced after completion of this study]. We also evaluated the accuracy of TS testing in people with varying CD4 counts.

## Materials and Methods

### Participant enrollment

Adults with HIV were enrolled in the parent DROP-TB study, a cohort diagnostic accuracy study of urine LAM and other TB testing methods (25). DROP-TB participants were enrolled at the time they presented for HIV care. Samples from a subset of the DROP-TB population (N=252) were tested in the TS sub-study. Between January 2022 and March 2024, adults (aged ≥18) living with HIV and initiating or re-initiating ART at outpatient clinics in uMkhanyakude district, KwaZulu-Natal, South Africa were enrolled regardless of self-reported TB symptoms. Consistent with inclusion criteria, participants self-reported no ART and no TB treatment in the preceding 90 days. After the first 100 participants were enrolled, a low TB case confirmation rate triggered a protocol modification designed to enrich the study population with persons with confirmed TB to reach enrollment targets.

Under the revised inclusion criteria, PWH were enrolled after receiving a positive sputum Cepheid Xpert MTB/RIF Ultra (Xpert Ultra) result, prior to initiating TB treatment. 21 participants enrolling under the revised inclusion criteria contributed samples to the TS sub-study. All participants provided sputum, urine, blood, and TS samples. Participants were eligible for evaluation in the TS sub-study if they provided at least one TS sample and had a final diagnosis of either pulmonary TB (sputum Xpert Ultra and/or sputum MGIT culture positive) or no TB (pulmonary or extrapulmonary).

### Ethics

All participants provided written informed consent. The study was approved by the University of Washington Institutional Review Board (Study 00006518), the University of KwaZulu-Natal Biomedical Research Ethics Committee (BREC/00001174/2020), and the London School of Hygiene and Tropical Medicine Ethics Committee (Ref. 21624). The study was also approved by the Health Research Committee of the KwaZulu-Natal Department of Health (NHRD KZ_202010_029).

### Sample collection

Research staff collected Medline DenTips pint-size foam swabs (Medline, MDS096202P) and Copan FLOQSwabs (Copan, 520CS01.BX) by brushing the dorsum of the tongue as described (12). Swabs were snapped or cut into 5-mL tubes (Globe Scientific, 6101S) or 2-mL tubes (Fisherbrand, 02-707-361), respectively. Samples were stored dry and frozen at – 80 °C by the research team at Africa Health Research Institute (AHRI) and later shipped on dry ice to the University of the Witwatersrand Diagnostic Innovation Hub (Wits DIH), Johannesburg, South Africa for processing and testing in September 2024. All swabs were collected prior to sputum collection. Sputum samples were transported to the National Health Laboratory Services (NHLS) lab at Hlabisa Hospital for testing with Xpert Ultra, and by courier to the AHRI clinical microbiology lab (Durban campus) for MGIT (Mycobacterium growth indicator tube) liquid culture.

The research nurse obtained a fingerstick capillary blood sample for hemoglobin A1C testing and up to 10 mL venous whole blood for research assays and storage. Research staff collected additional blood for routine HIV care initiation labs (CD4 T-cell count, creatinine, hemoglobin, hepatitis B surface antigen) as part of the same venipuncture. Blood samples were tested at the Hlabisa NHLS facility. CD4 T-cell counts were measured from whole blood as part of routine care according to South African national guidelines for HIV treatment and monitoring. Laboratory values were abstracted from the NHLS LabTrak system.

Study participants were provided with urine collection cups and requested to provide a 50-200 mL urine sample. Urine samples were transported to the AHRI lab in Somkhele for Abbott Determine™ TB LAM Ag testing (LF-LAM) following manufacturer protocols. Results were recorded in the study REDCap database.

### Case definition

Sputa were tested by Xpert Ultra by NHLS and MGIT culture in the AHRI laboratory to generate the extended MRS (eMRS). A positive TB eMRS result was defined as at least one of 1) a positive or trace sputum Xpert Ultra result or 2) a positive sputum MGIT culture result. Although Xpert Ultra trace results are not considered confirmation of TB disease in some settings, participants with trace results in this study were assessed as having TB by non-study clinicians and placed on TB treatment. A negative TB eMRS was defined as at least one negative result and no positive results from sputum Xpert Ultra and TB culture, typically seen as negative results for both sputum Xpert Ultra *and* TB culture. If one of the two microbiologic reference tests was negative and the other was indeterminant, not performed, or contaminated, the eMRS was considered negative.

### TS sub-study sample selection

A subset of the population was identified for testing by SSMaC in a 1:3 ratio of people with a positive vs. negative eMRS result, including all people with a positive eMRS. This subset list of participant identification numbers was generated by an investigator who was unblinded to the eMRS status of the participants, but who was not involved in the SSMaC testing. TB-negative controls were matched to cases based on the date of sample collection. All investigators involved in swab testing remained blinded to matching strategy, clinical characteristics, eMRS, and LF-LAM results of participants until swab testing results were confirmed. Included and excluded samples are illustrated in Figure 1. A preliminary pilot set of N = 40 samples were analyzed and subsequently unblinded to eMRS prior to testing the rest of the sample set. This was to confirm assay performance. All clinical specimens (cases and controls) collected per protocol with the intended foam product stored in the intended tubes were analyzed in this arm. Some samples (N = 56) were collected or stored off-protocol and were excluded from analysis **(Figure 1)**.

**Figure 1.**
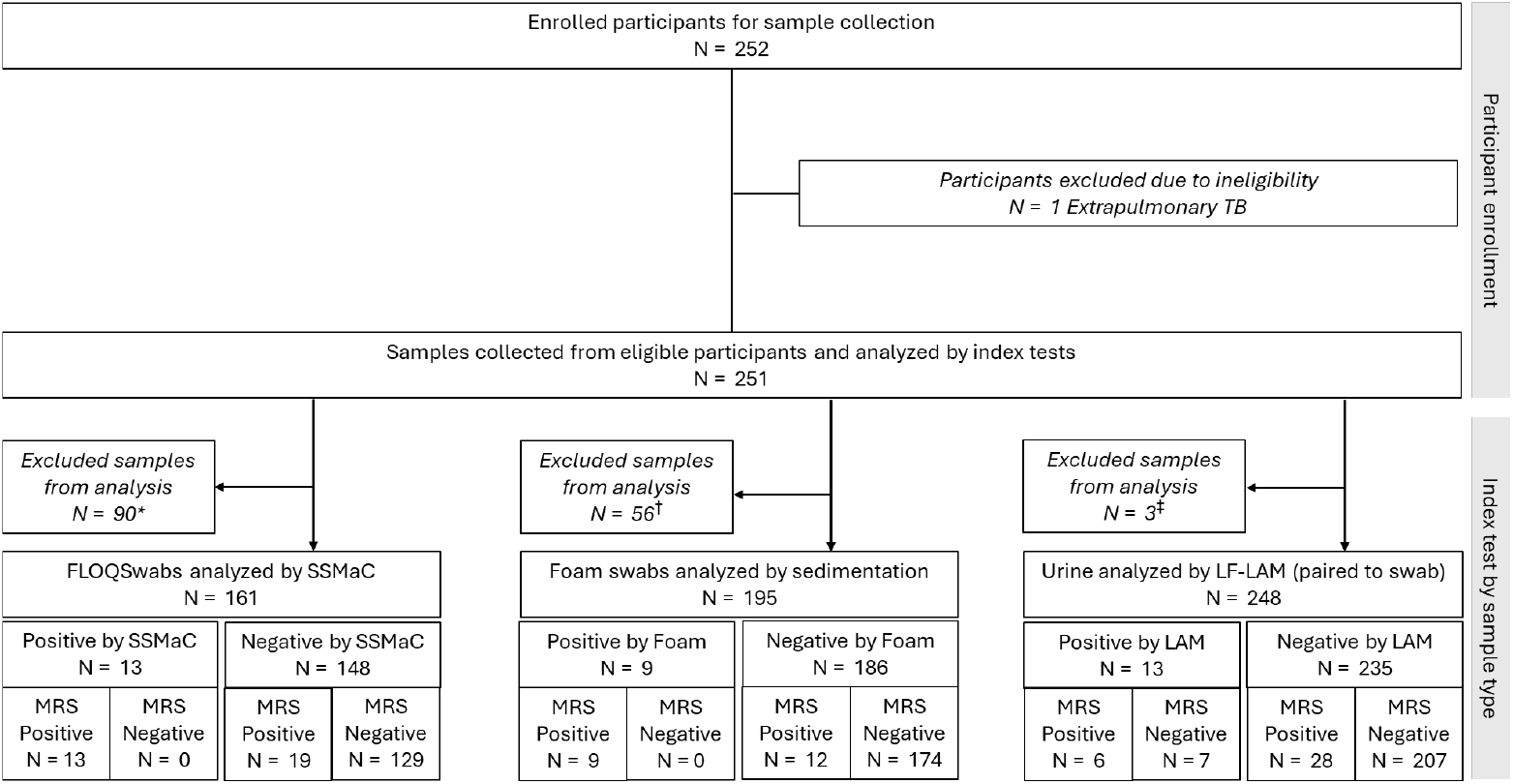
STARD diagram showing the flow of participant inclusion, sample testing, and results. Exclusions were for the following reasons: *Swabs not selected through subset matching algorithm; ^†^Non-protocol, incompatible swab type or storage used in sample collection; ^‡^data unavailable for analyses.

### FLOQSwab-SSMaC processing and qPCR

FLOQSwabs were analyzed as previously described (12) with the following modifications: A thermomixer (Eppendorf, 2231001127) was used for incubations at elevated temperatures (55 °C, 100 °C and 95 °C); the 1 hour hybridization at ambient temperature was done using a sample rotator (Benchmark Scientific, SKU:RS3020) on speed 30; and qPCR was done on a QuantStudio 7 Flex Real-Time PCR system (Applied Biosystems, 4485701). All runs included a standard curve of 1:10 dilution series starting with 5 ng H37Rv DNA (ATCC) and water no-template controls. Results were exported with a normalized threshold of 750,000 and analyzed in Microsoft Excel. Results were determined to be positive if <40 Cq. For Cq values >40 results were considered negative.

### Foam-sedimentation processing and qPCR

Foam swab samples were analyzed as previously described (12) with the following modifications: Pelleted samples were mechanically lysed on a Vortex-Genie 2 with an adapter (SI-H524, Horizontal Microtube Holder (24 tubes), Scientific Industries Inc, NY, USA), shaking horizontally at maximum speed for 20 min; the IS*1081* probe was used with a FAM fluorophore, as in the SSMaC assay; and qPCR was run on a QuantStudio 7 Flex. All runs included a standard curve and water NTCs. Results were exported with a normalized threshold of 300,000 and analyzed in Microsoft Excel.

### Extraction control preparation

FLOQSwabs were self-collected by healthy volunteers in Seattle, WA, USA, and snapped into 2-mL screw-capped tubes. Samples were spiked with 10 µL of cultured MTB H37Rv cells diluted in PBS for use as positive extraction controls. Negative extraction controls were not spiked and were frozen immediately after collection. Foam swab controls were spiked with 40 µL of the same diluted cell cultures. Negative extraction controls for the foam arm of the study were plain TE buffer samples without tongue material present. All positive extraction controls were transported on ice to the lab for processing alongside clinical samples by the department arranged courier and frozen that afternoon. Cells were prepared in advance, grown in MGIT, and then diluted to 1 McFarland unit. These stocks were frozen and later diluted for spiking.

### Diagnostic accuracy calculations

Sensitivity and specificity values were compared using two-population Z tests of proportion.

## Results

Samples from 251 participants were evaluated in the TS study. The study population was 56% female with a median age of 34 years (Table 1). The median CD4 count was 347 cells/mm^3^, with a quarter of the population less than 200 cells/mm^3^, indicating advanced HIV disease status.

**Table 1.** Baseline demographics and clinical characteristics.

| Characteristic | All N=251 (%) | eMRS-positive participants among total N=32 (%) (Trace = positive) | eMRS-negative participants among total N=219 (%) (Trace = positive) |
| --- | --- | --- | --- |
| Male | 110 (44) | 22 (69) | 88 (40) |
| Female | 141 (56) | 10 (31) | 130 (60) |
| Age (median, IQR) | 34 (27-40) | 39 (34-50) | 33 (27-39) |
| CD4 (median, IQR) | 347 (184-550) | 211 (106-512) | 363 (199-555) |
| BMI (median, IQR) | 22.9 (20.3-27.2) | 20.2 (18.7-22.7) | 23.3 (20.9-27.6) |
| TB Symptoms (4QSS+)* | 168 (67) | 30 (94) | 138 (63) |
| No TB Symptoms (4QSS-)* | 83 (33) | 2 (6) | 81 (37) |
| Prior TB | 40 (16) | 9 (28) | 31 (14) |

\*4QSS+, yes to one or more 4-question WHO symptoms, 4QSS-, no to all 4-question WHO symptoms

Approximately one third of the study population did not report TB symptoms in the 4-question WHO symptom screen (4QSS) and were categorized as “No TB Symptoms”. Most participants (67%) presented with at least one self-reported symptom. A prior TB diagnosis was noted for a small portion of the population (16%).

The study population was 13% (32/251) sputum eMRS positive. The 32 eMRS positive participants included 14 who were positive by Xpert Ultra only (including trace Xpert Ultra), and 1 who was positive by culture only (**Table 2**). This group also included 21 eMRS positive participants who were enrolled under the revised criteria established to enrich for people with confirmed TB.

**Table 2.** TB eMRS results from sputum.

| TB results | n/N (%) |
| --- | --- |
| Total TB eMRS negative | 219/251 (87) |
| Total TB eMRS positive | 32/251 (13) |
| Total TB culture positive | 18/251 (7) |
| TB Xpert Ultra positive (includes trace) | 31/251 (12) |
| TB-enriched population | 21/251 (8) |

**Table 3** shows the sensitivity and specificity of each of the two swab methods relative to eMRS. Foam-sedimentation and FLOQSwab-SSMaC performed similarly to each other, with 47% and 43% sensitivity, respectively (*p* = 0.7948). Sensitivity of both methods was higher when trace Xpert Ultra results were classified as TB negative. Specificity was ≥99% by both swab methods. Both methods were significantly more sensitive than LAM, regardless of whether eMRS included Xpert Ultra trace as positive (two-tailed z-scores ranging from 0.01 to 0.03).

**Table 3.** Foam swab-sedimentation, FLOQSwab-SSMaC, and LF-LAM sensitivity and specificity (with and without sputum trace results considered as eMRS positive) and Z test results.

|  | Foam-sedimentation N=195 |  | FLOQSwab-SSMaC N=161 |  | LF-LAM N=248 |  |
| --- | --- | --- | --- | --- | --- | --- |
| Reference Standard | eMRS<br>Trace =<br>Positive | eMRS<br>Trace =<br>Negative | eMRS<br>Trace =<br>Positive | eMRS<br>Trace =<br>Negative | eMRS<br>Trace =<br>Positive | eMRS<br>Trace =<br>Negative |
| **Sensitivity n/N** | 9/19 | 8/16 | 13/30 | 13/25 | 6/32 | 5/27 |
| **Sensitivity** | 47% | 50% | 43% | 52% | 19% | 19% |
| **(95% CI)** | (24-71%) | (25-75%) | (25-63%) | (31-72%) | (7-36%) | (6-38%) |
| *P value compared to FLOQSwab-SSMaC\** | 0.78 | 0.90 |  |  | 0.04** | 0.01** |
| *P value compared to foam-sedimentation\** |  |  | 0.78 | 0.90 | 0.03** | 0.03** |
| **Specificity n/N** | 176/176 | 178/179 | 131/131 | 136/136 | 209/216 | 213/221 |
| **Specificity** | 100% | 99% | 100% | 100% | 97% | 96% |
| **(95% CI)** | (98-100%) | (97-100%) | (97-100%) | (97-100%) | (93-99%) | (93-98%) |
| *P value compared to SSMaC\** | NA*** | 0.38 |  |  | 0.04** | 0.03** |
| *P value compared to foam swab\** |  |  | NA*** | 0.38 | 0.02** | 0.04** |

\*Two population Z test, two tailed
\*\*Significant at p < 0.05
\*\*\*Not applicable; the numbers are identical

We evaluated diagnostic sensitivity and specificity of TS (both foam-sedimentation and FLOQSwab-SSMaC) and LF-LAM stratified by CD4 count (**Table 4**). Foam-sedimentation and FLOQSwab-SSMaC were more sensitive among participants with CD4 counts <200, compared to those with CD4 ≥200. Overall, TS was more sensitive than urine LAM across CD4 strata. However, the study was not sufficiently powered to confirm differences across strata and confidence intervals for all estimates overlapped (**Table 4**).

**Table 4.** Foam, SSMaC, and LF-LAM sensitivity and specificity stratified by CD4 count (≥ or < 200) eMRS.

|  | Foam<br>swab<br>CD4≥200<br>N = 141 | Foam<br>swab<br>CD4<200<br>N = 51 | SSMaC<br>CD4≥200<br>N = 115 | SSMaC<br>CD4<200<br>N = 43 | LF-LAM<br>CD4≥200<br>N = 174 | LF-LAM<br>CD4<200<br>N = 69 |
| --- | --- | --- | --- | --- | --- | --- |
| **Sensitivity (n/N)** | 3/9 | 6/10 | 6/16 | 7/14 | 2/17 | 4/15 |
| **Sensitivity**<br>**(95% CI)** | 33%<br>(8-70%) | 60%<br>(26-88%) | 38%<br>(15-65%) | 50%<br>(23-77%) | 11%<br>(1-36%) | 27%<br>(8-55%) |
| **Specificity (n/N)** | 132/132 | 41/41 | 99/99 | 29/29 | 153/157 | 51/54 |
| **Specificity**<br>**(95% CI)** | 100%<br>(97-100%) | 100%<br>(91-100%) | 100%<br>(96-100%) | 100%<br>(88-100%) | 98%<br>(94-99%) | 94%<br>(85-99%) |

The average number of symptoms (0-4) were not statistically different in participants correctly identified (2 ± 1.1) and missed (2 ± 1.2) by FLOQSwab-SSMaC. Similarly, foam-sedimentation detected versus missed averaged 2 ± 0.8 and 2 ± 1.0 average symptom score, respectively.

Test agreement among participants who were tested with all 3 index tests (SSMaC, foam swab, and urine LAM) is summarized in a Venn diagram in **Supplemental Figure 1**. Most sputum-positive participants with positive swabs were positive by both swab methods (FLOQSwab-SSMaC and foam-sedimentation). However, two eMRS-positive participants (including one who was sputum trace-positive) were positive only by foam-sedimentation, and one eMRS-positive participant was positive only by FLOQSwab-SSMaC. The urine LAM test results aligned with the two swab methods in half (N = 4) of the 8 cases detected by urine LAM. Seven eMRS-positive participants were not detected by any of the three index tests.

## Discussion

Two tongue swab-based TB detection methods were evaluated in a population of 251 South African PWH irrespective of TB symptoms. FLOQSwab-SSMaC (N=161) and foam-sedimentation (N=195) performed similarly, achieving 43% to 47% sensitivity, respectively, relative to eMRS. Categorizing sputum “trace” results as negative in the eMRS consistently the apparent sensitivities of both TS methods (52% and 50%, respectively). In addition, LAM was detected in corresponding urine samples using a commercial LF-LAM (N=248), with an overall sensitivity of 19% compared to the eMRS. Specificity relative to sputum eMRS was high at ≥99% for both TS methods and 97% for urine LAM.

It has been well demonstrated that the sensitivity of TS testing for TB declines in people with low bacillary loads, for example patients with semi-quantitative sputum Xpert Ultra grades in the “trace” or “very low” range (12, 13, 15, 18, 19, 26). Therefore, it was not surprising to find modest sensitivity of TS testing within this active case-finding population, in which participants were not actively seeking TB care and may have had low bacillary loads. Despite this limitation, the ease-of-use of TS collection relative to sputum collection, especially in non-clinical settings and/or among PWH, may facilitate TB case finding in contexts such as this. While better sensitivity would be desirable, the sensitivity seen here (about half of eMRS-positive TB cases were detected), could make TS testing useful in settings where sputum collection is not practical.

Recently, members of our study team reported that the sensitivity of TS testing was lower in an asymptomatic, community-based TB screening population than in a symptomatic clinical population (20). The current study was not powered to make a similar comparison. However, the average number of TB symptoms was not significantly different in participants with TB correctly detected vs. missed by either TS method.

The three non-sputum TB testing methods were more sensitive among people with CD4 counts <200 cells/µl than among people with CD4 counts ≥200 cells/µl. Such observations are well documented with urine LAM testing (27-29). However, to our knowledge this is the first report of a similar observation in TS testing. Interestingly, some previous studies reported a trend toward lower sensitivity of TS testing in PWH relative to people without HIV (12, 14). Our results suggest that this trend may not be related to immune status, and instead may be explained by other demographic or patient care-related factors. Notably, one previous study reported higher sensitivity of oral (buccal) swab testing in PWH compared to people without HIV (30).

In this study population, all participants with a positive sputum Xpert Ultra result, including trace, were recommended to start anti-TB treatment. Therefore, participants with sputum Xpert Ultra trace results were included in our diagnostic accuracy calculations. However, in some settings, people with Xpert Ultra trace results are not considered to have confirmed TB (31, 32). Therefore, we calculated accuracy in two ways, with and without the inclusion of Xpert Ultra trace participants among the confirmed TB cases. Removal of trace from the MRS definition resulted in apparently increased sensitivities observed in the index tests. In this study population, sputum Xpert Ultra semi-quantitative results were not available, other than “trace”. Future studies evaluating TS methods should consistently collect and report semiquantitative grade, and should be expanded to enable more extensive stratification based on symptoms and other metrics of disease severity.

This study had several additional limitations. The population was relatively young with a median age of 34, and 56% of participants were female. Globally, more TB occurs in older males, so some of these findings may not be generalizable. However, the population of PWH who are initiating ART is representative of an important potential use case for TS. Another limitation was that the enrolled participants were all able to spontaneously expectorate sputum with coaching from study personnel.

Sputum-scarce, or persons unable to spontaneously expectorate sputum, were not represented, and it is estimated that sputum-scarce persons comprise nearly 25% of all adults and adolescents being evaluated for TB (33). It will be useful to evaluate the performance of TS in sputum-scarce persons. This may be one of the most impactful applications of the TS method, since even modest sensitivity is an improvement over zero sensitivity when relying on sputum-based diagnostics for a sputum-scarce person. An additional limitation is that the number of confirmed TB-positive participants was small in this study population. Finally, new nPOC TS testing platforms (19), which were recommended recently by WHO (21), were not available for evaluation at the time of this study.

In summary, we evaluated the ability of TS and urine LAM to detect TB disease among non-TB care-seeking PWH. TS methods combined with qPCR-based testing detected about half of TB cases within this population, and were more sensitive than urine LAM testing, including among people with low CD4 counts. By helping to facilitate TB screenings in populations such as this one, TB methodologies could improve patient care and help to interrupt TB transmission among PWH and other at-risk populations.

## Data Availability

All data produced in the present study are available upon reasonable request to the authors.

## Acknowledgements

We are grateful to the people of uMkhanyakude for participating in our study and to the uMkhanyakude District Department of Health and clinic staff for their involvement. The study was funded by NIH/NIAID grant K23 AI140918 to AES, with additional support from R21 AI185407 to GAC.

