## Supplemental information for "Tongue swab-based Targeted Universal Tuberculosis Testing in people living with HIV in KwaZulu-Natal, South Africa"

**SUPPLEMENTAL FILES**

**Supplemental Figure 1. Venn Diagram of all method results N = 102.** Numbers within the circles represent positive results by each index test and reference test method. Participant with sputum "trace" results in the reference test are shown separately within blue circles.

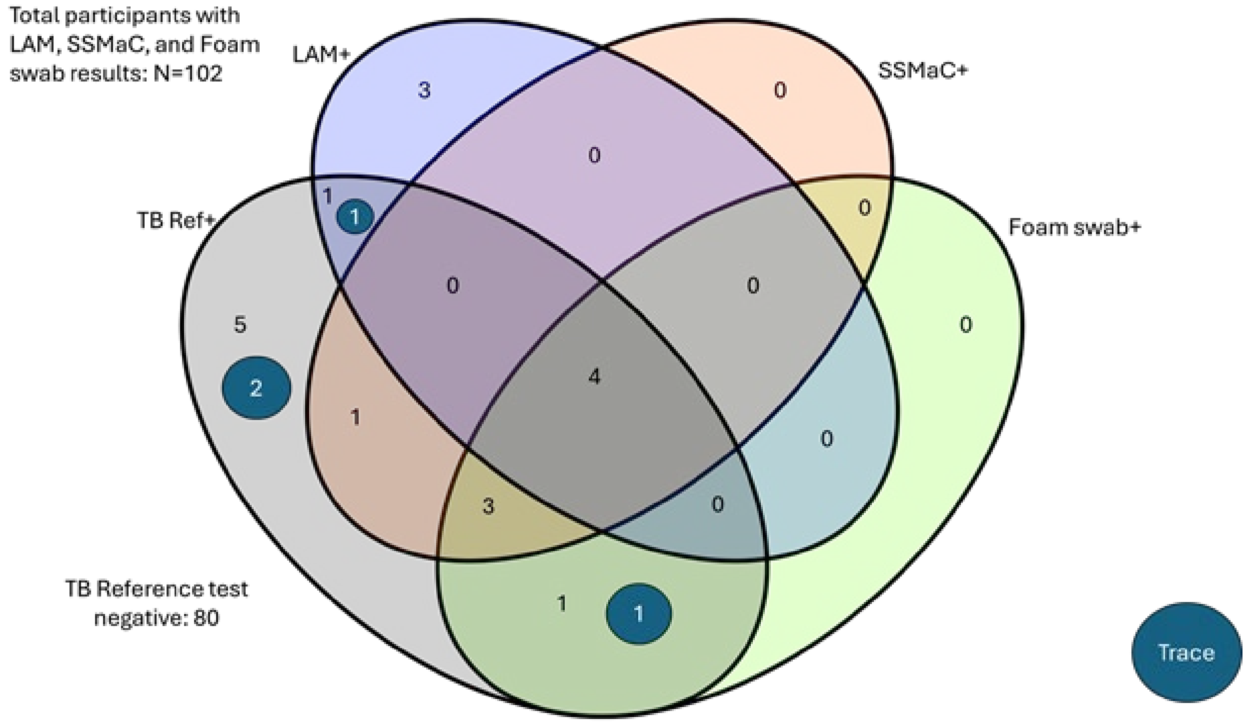
